# Putting (Big) Data in Action: Saving Lives with Countrywide Population Movement Monitoring Using Mobile Devices during the COVID‐19 Crisis

**DOI:** 10.1101/2020.09.21.20194019

**Authors:** Miklos Szocska, Peter Pollner, Istvan Schiszler, Tamas Joo, Tamas Palicz, Martin McKee, Magyar Telekom Nyrt., Telenor Magyarorszag Zrt., Adam Sohonyai, Jozsef Szoke, Adam Toth, Peter Gaal

## Abstract

Many countries have implemented strict social distancing measures in the hope of reducing transmission of SARS-CoV-2 but the effectiveness of these measures is determined by the willingness of populations to comply with restrictions. Consequently, a system of monitoring population movement using existing data sources can inform those making decisions about policy responses to the COVID-19 pandemic.

We describe a collaboration with all 3 major domestic telecommunication companies in Hungary to use aggregated anonymous mobile phone usage data to calculate two indices for assessing the effect of movement restrictions: a “mobility-index” and a “stay-at-home (or resting) index”. The strengths and weaknesses of this approach are compared with the smartphone-based, COVID-19 Community Mobility Reports from Google.

Data generated by mobile phones have long been identified as a potential means to analyse mass population movement, but its operationalisation raises several technical questions, such as making sense of Call Detail Records, collation of data from different mobile network providers, and personal data protection concerns. The method described here addresses these issues and offers an effective and inexpensive tool to monitor the impact of social distancing measures, achieving high levels of accuracy and resolution. Especially in populations where uptake of smartphones is modest, this method has certain advantages over app-based solutions, with greater population coverage, but it is not an alternative to smartphone-based solutions used for contact tracing and quarantine monitoring. We believe that this method can easily be adapted by other countries.

## Introduction

The COVID-19 pandemic, which began at the end of 2019, prompted governments of virtually all countries in the world to implement strict social distancing measures on a virtually unprecedented scale.^1–8^ They have proven themselves very effective in slowing down and, in many countries, bringing under control the infection, but with severe economic costs and social sacrifices.^9–12^ Indeed, they are not without adverse health consequences, acting through increases in mental illness and deferred health care, for example. Thus, the social and the economic consequences of the pandemic and of the measures to address it are inextricably intertwined.^13^ Consequently, governments must find a delicate balance between taking measures to bring the pandemic under control as rapidly as possible while minimising social and economic costs.

Hungary is among the fortunate countries in Europe during the pandemic.^14^ It was one of the first EU member states to declare a state of emergency, accompanied by strict social distancing measures.^15^ The success of such measures in interrupting the exponential growth of cases does, however, depend on how well people comply with the regulations.

The use of data from mobile phones to analyse physical mobility in a population is not new,^16–19^ and the location of mobile devices, as a proxy for the users’ geographical presence, has long been used for commercial purposes, such as for tailored advertising, as well as to inform subscribers about their daily activities, such as numbers of steps taken.^20^ The opportunity that mobile phones and associated apps offer in combating the pandemic has already been recognised,^21^ with applications such as the Chinese coronavirus “close contact detector” app, which brings together routine data on travel, case reports, and users’ mobile phone location information to detect possible contacts with infected people,^22^ or the South Korean official mobile phone apps, which are used similarly for agile contact tracing, but also as a means to monitor people in home quarantine.^23^ There are also accounts of using mobile phone data to assess the impact of social distancing measures from several EU countries, as well as China, Chile, the US, and the UK.^21^ While the pooling of personal information, such as GPS data from mobile phones, car navigation systems, credit card use, or security camera footage, is fraught with legal and ethical questions, digital contact tracing has been suggested as a way to ease social distancing measures while keeping the pandemic at bay.^24^ While recognising the ethical and legal considerations in using such data, the immense potential of the methodology justifies the attention given to it, while international collaborations offer scope to bring together the still somewhat limited expertise that exists, especially in smaller countries.^21^

In this paper, we present an example from Hungary of how health data science, using data generated by mobile phone usage, could assist the management of the COVID-19 pandemic. In particular, we describe the use of anonymized aggregate cellular data for monitoring of the effectiveness of social distancing measures. Our purpose is to share our experiences with other countries interested in replicating this work.

## Monitoring the impact of social distancing measures in Hungary

Italy was the first European country to experience COVID-19, with scenes of struggling hospitals acting as a wake-up call to those who had initially downplayed the threat from the disease.^25 26^ As the crisis in Italy unfolded, other European countries started to act, implementing severe social distancing and isolation measures, in some cases very rapidly.^5^ In Hungary, the first two known COVID-19 cases were confirmed on 4 March. Visits to hospitals and long-term care facilities were banned on 8 March and a state of emergency was declared by the government on 11 March, restricting public gatherings and requiring universities to switch from face-to-face teaching to distance learning.^27 28^ This was followed by school closures, announced on 13 March and effective from 16 March, when all public gatherings were banned and severe restrictions were imposed on retail trade and personal services. Eventually a relatively strict curfew was implemented on 28 March.^29–31^ However, this was not a total lockdown, with outdoor recreational activities still permitted, giving rise to some crowding around popular tourist destinations, but mayors were authorised to implement local restrictions to prevent this.^32^ Travel and entry restrictions followed the same pattern, starting with suspending entry only from hard hit countries such as China, Iran, Italy and South Korea on 11 March, adding Israel on 14 March, with full border closures on 16 March.^28 33 34^ The speed with which it acted places Hungary among the fastest responding countries in Europe. For instance, measured as the delay between confirmation of the first COVID-19 cases until implementation of a nation-wide school closure and switch to distance learning, Hungary was the eighth fastest in the EU-27 and the UK.^5^

Effective decision making in a crisis is greatly facilitated by access to relevant information. The Hungarian Ministry for Innovation and Technology (which has been authorised by the government to access routinely available data to support the management of the pandemic)^35^ in collaboration with data experts and researchers, established a Digital Health and Data Utilisation Team (DHDUT) on 22 March, tasked with identifying relevant data that could inform the design and implementation of responses to the pandemic. Using raw data as diverse as medical information from the Hungarian eHealth cloud, financing data from the National Health Insurance Fund Management, epidemiological data from the National Public Health Center and Call Detail Records (CDR) from the domestic telecommunication companies, the experts quickly assembled a dashboard reporting both standard indicators and graphs, such as numbers of new and cumulative cases, deaths, and recoveries and their age, gender, and geographical distribution, but also developed new indicators, for instance for monitoring the effectiveness of social distancing measures. This dashboard now serves as the main management information system for those members of the government, involved in the day to day pandemic decision making. The aim of the work reported here was to develop a measure to monitor the effectiveness of social distancing measures, a so-called mobility (and stay-at-home) index.

## Methods

We sought to develop an index to track changes in population movement during the imposition of social distancing measures, comparing results from before and after the implementation of restrictions. We used geolocation data generated by the use of mobile phones. Mobile network providers locate users whenever they use their phone to initiate communication, whose parameters are automatically stored in so-called Call Detail Records (CDR). This communication can be of several types: calling another client, sending an SMS, or starting a new data transfer session during use of the Internet. The recorded location data for each communication session are the location of the cellular base station (also referred to as cellular tower or mast), which handled the transmission request. When a mobile phone is connected to more than one mast, this is the closest one with the best signal and available capacity.

Hungary has three main companies providing mobile telecommunications for the general public, with market shares of 44.8%, 27.2%, and 26.7%, as of the end of 2019.^36^ There is one new company with its own network infrastructure and few other companies, which provide services using the network infrastructure of the 3 big companies, but their market share is negligible, at 1.3% for calls and 2.3% for internet service provision. The three telecommunications companies provided daily CDR data, starting on 15 December 2019, and collaborated with the research team to develop the methodology and advise on how to interpret the raw data for the purpose of our analysis.

Given the need to respect the anonymity of clients and to remain compliant with the European Union General Data Protection Regulation (GDPR), each provider aggregated data at the level of settlements, the lowest level of public administration in Hungary. In 2019, there were 3,155 settlements with an area ranging from 1 km^2^ to 488 km^2^ and a population from 10 to 201,432, except for the capital, Budapest, which covers an area of 525 km^2^ with a population of more than 1.7 million, 18% of the total population of Hungary.^37^ Since Budapest is divided into 23 districts with own local governments, each district was treated as a separate settlement. Close to 53% of the population lives in one of the other 345 towns, while only 3% lives in small villages with less than 500 inhabitants. A phone was defined as changing location if it was registered in two or more settlements in a 24 hour period and non-mobile if it remained in only one. The mobile phone geolocation data places a phone within the land area served by the transmitting cellular tower, whose radius can be as large as 35 kilometres, but the actual geographical resolution that can be achieved depends on the density of cell towers, which is much higher in densely populated areas, because the capacity of masts to handle simultaneous communication sessions are limited. In urban areas, this can be as low as a few hundred metres, while in rural areas masts are generally spaced at least a few kilometres apart. Each mast was assigned to a settlement by the telecommunications companies, which ensured an unequivocal matching of CDR location data with settlements.

The aggregated datasets from the three providers in Hungary were then merged by the DHDUT for each settlement. These figures were used to create 2 indices, a “mobile” and a “stay-at-home” index. Since the mobility of the Hungarian population fluctuates over the course of a week, a simple sum of SIM cards in each category would be misleading for our purpose, so the values were normalised in relation to the distribution by day of week in a reference period, in this case February 2020. Hence, the relative-mobility and the relative-stay-at-home index are calculated as the ratio of the device counts during a given day in the pandemic period to the average value for the corresponding weekdays in the reference period. Thus, the relative-mobility index on 14 April is calculated as 100·*c*0414*/CTuesday*100, where *c*_0414_ is the count of devices that have changed their location on 14 April (Tuesday) and *CTuesday* is the average device-count of Tuesdays of the reference month. National holidays were normalised as Sundays.

Finally, we compared our results with those generated using Google mobility data. Although both use data from mobile phones, they differ in that the Google data track individuals continuously, either via satellites, cellular base stations or both, but only as long as their location function is turned on, whereas CDR automatically register the location data, but only when someone interacts with the telephony provider. Further, the former only captures mobility of smartphones while the latter captures all mobile phones. Hence, one would expect that the measures would differ, but it is not clear how much and in what way.

## Results

Trends in numbers of devices, prior to adjustment for day of week, are shown in Fig. 1. A clear weekly pattern is visible, with a substantial reduction in movement at weekends, with the reduction most marked on Sundays. Higher activity on working days with a relatively strong maximum on Fridays can be observed as well. In contrast, the resting dataset is not so volatile, showing only a decent maximum on Sundays and a minimum on Fridays.

**Figure 1.**
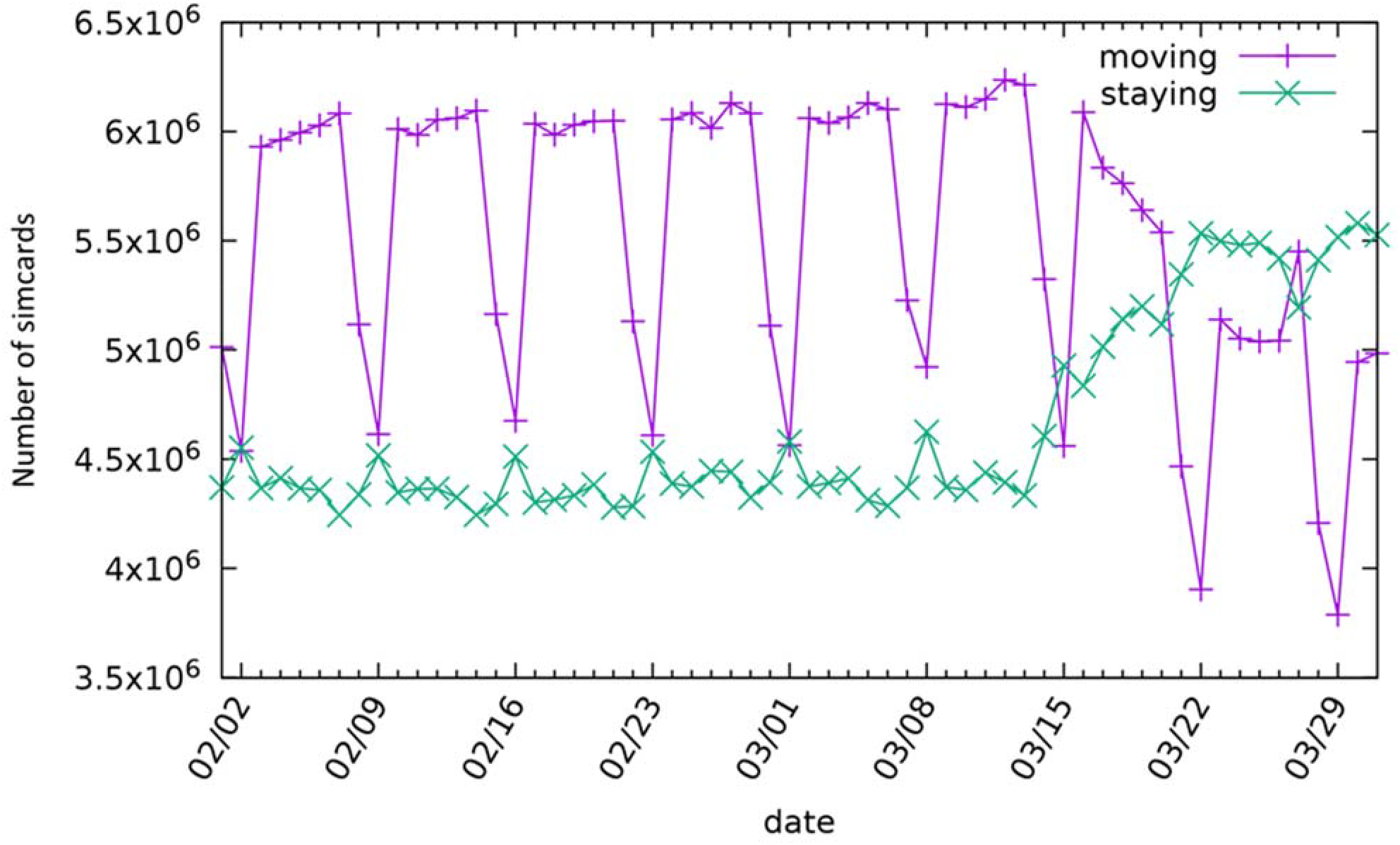
Daily device activity in Hungary for February and March of 2020.

After normalising to the reference values, the weekly periodicity has smoothed as shown in Fig. 2. The trend of the two indices can be considered to reflect what happened when the government imposed restrictions on movement. As noted above, the first significant measures were introduced on 11 March and continued with the closure of schools on 16 March. This was associated with a clear drop in mobility, which is intuitive as parents had to remain at home to care for their children. Two weeks later, the staying at home regulation was announced, which was a lighter version of curfews elsewhere. As Fig. 2 shows, both mobility and resting indices remained relatively stable, except for a discrete peak associated with announcement of the regulation. On this day, many members of the public rushed to supermarkets and pharmacies to prepare themselves for the forthcoming restrictions on mobility. Fig. 2 also shows the impact of the gradual loosening of movement restrictions, which, in Hungary, started on 30 April, with the state of emergency being abolished on 18 June.

**Figure 2.**
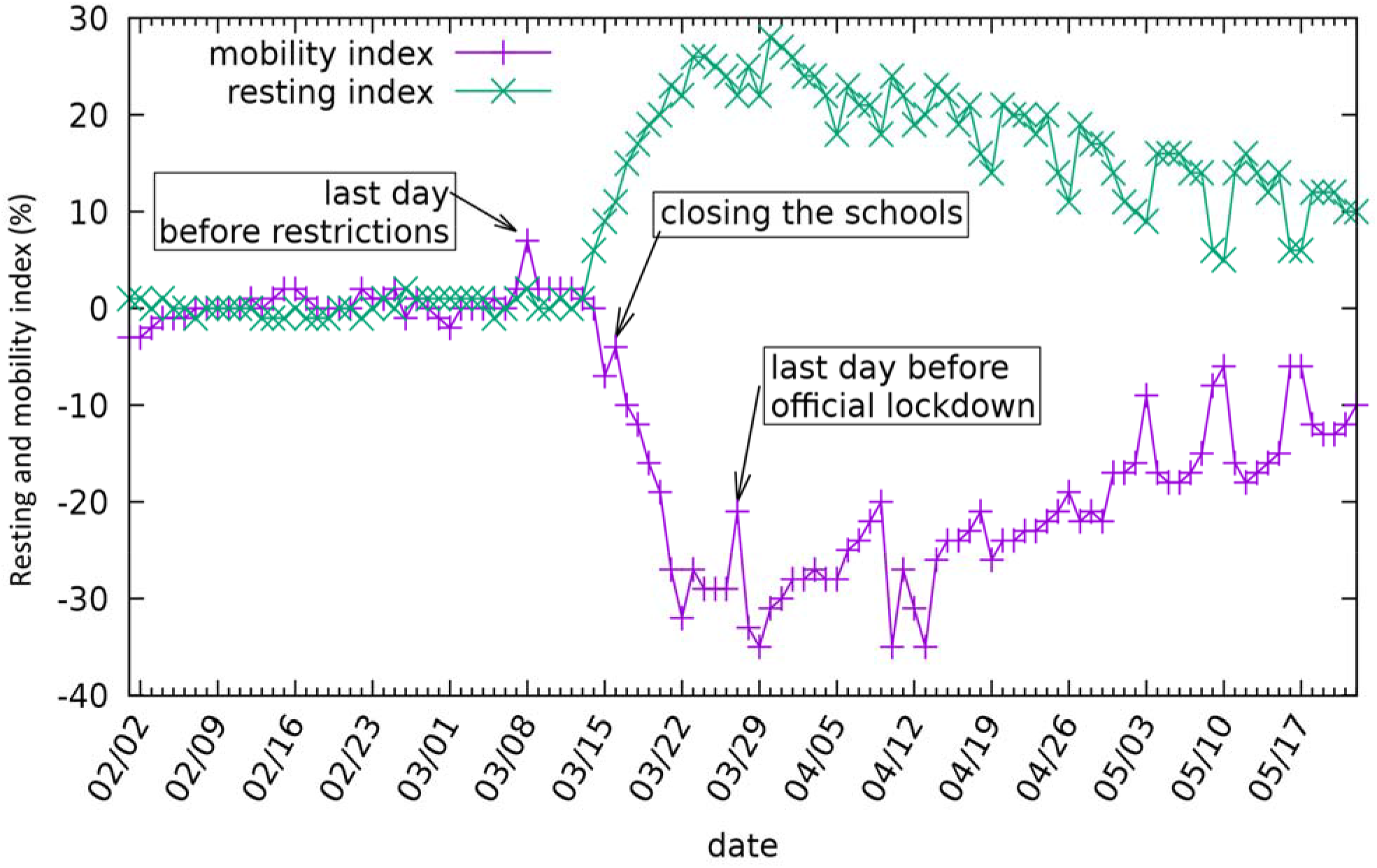
The mobility and stay-at-home (resting) indices for Hungary, showing the civil response to government measures

Moving to the local level, the data are sufficiently granular to track changes at the level of individual settlements in smaller geographical areas. This is illustrated in Fig. 3, showing the change in the mobility index at the level of settlements in 3 Hungarian counties (Somogy, Zala and Veszprém) around Lake Balaton on 5 April. At the end of March, rumours started to spread in various media that people with a weekend house near Lake Balaton (one of the most popular domestic tourist destinations during the Summer), decided to move from Budapest and other cities in large numbers with the intention of staying in quarantine there. Initially this story was supported only by anecdotal evidence, however the closer examination of the mobility index at the local level confirmed the problem. The majority of the settlements close to Lake Balaton are coloured dark in Fig. 3, signalling an anomalous increase of at least 50%, but in certain cases as high as 800% in the mobility index, and similar intensification of movement could be found in other popular hiking places around the country, such as in the vicinity of Budapest. In response, mayors were authorised to impose local movement restrictions in addition to the national measures if they felt necessary.

**Figure 3.**
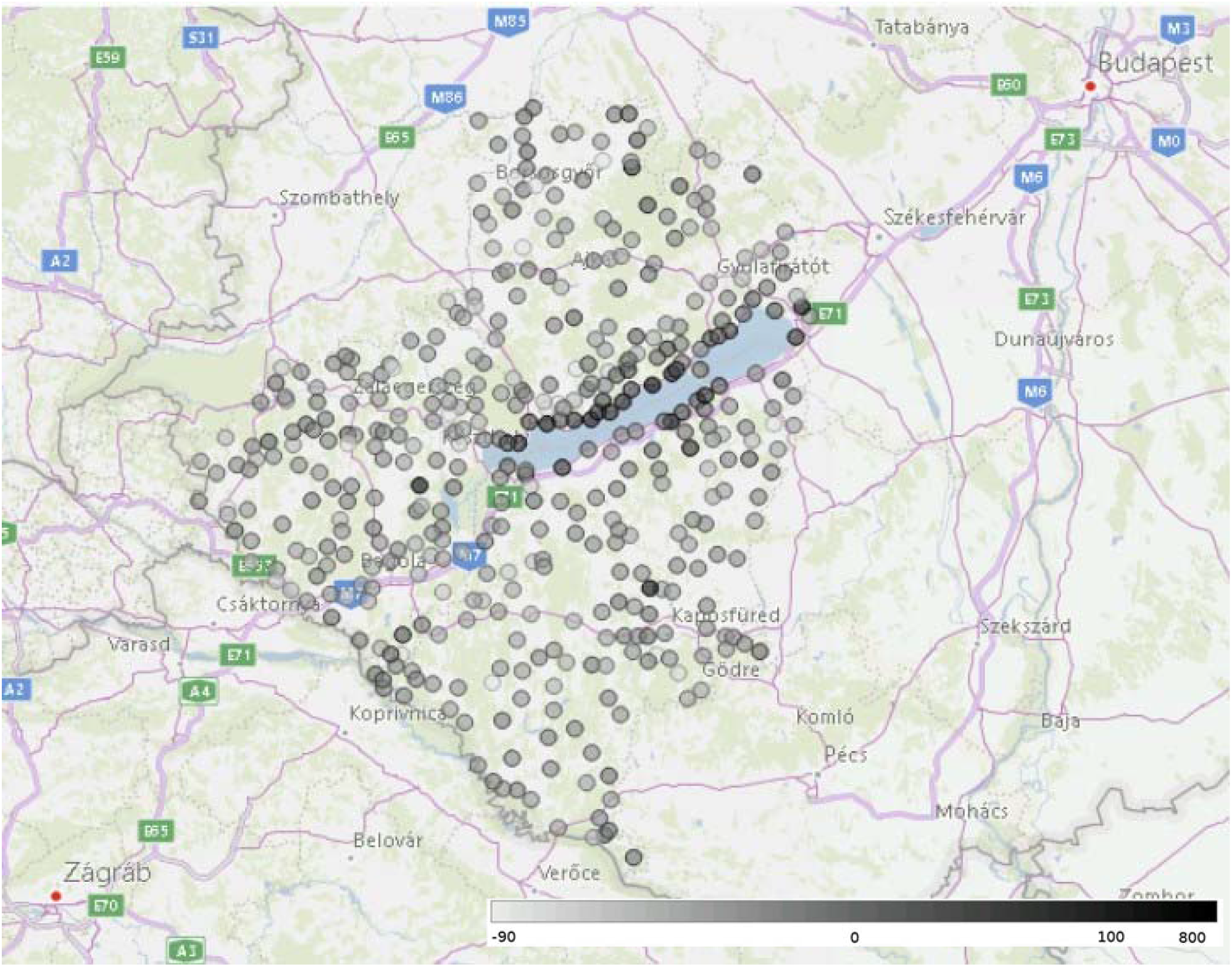
The changes in the mobility index in the settlements (each represented by a shaded circle) around one favourite holiday place in Hungary, the lake Balaton on 5 April 2020. Lighter grey colouring represents a decrease in mobility compared to the reference day, while darker grey colouring represents an increase.

As noted above, Google and other global technology companies use smart phone generated data to track the location of users. We looked at the COVID-19 Community Mobility Reports from Google in detail.^38^ Its main features are essentially the same as features of other smartphone-based methods, when a specific smartphone application is developed to track the movement of those, who download and install this. While, as noted above, these are usually applied to track individuals or smaller population groups for the purpose of monitoring targeted interventions, such as contact tracing or quarantine monitoring, it is technically possible to use them for the monitoring of large scale population movement. As such, they are an alternative to CDR-based methods. The comparison with the mobility reports of Google, therefore, serves both as an external reference point for validation, but also to set out their relative advantages and disadvantages when policymakers must decide on solutions to be deployed in their countries.

In Fig. 4a we compare the CDR-based resting index calculated between 16 February and 17 May and the residential index of Google, based on similar aggregated and anonymized data used to show popular times for places in Google Maps. The curves follow very similar patterns, e.g., the effect of the curfew announced on 11 March is manifested in a large increase of both indices. In Fig. 4b we show a scatter plot of the daily CDR-based resting (staying-at-home) index as a function of the daily residential index from Google. Most of the points are close to the diagonal line that would correspond to a perfect fit. However, during the weekends our CDR-based index takes a larger value compared to the Google index (points deviate from the diagonal to the left), whereas on national holidays the opposite happens and the Google index is larger. The likely reason behind this behaviour is the different reference baseline, e.g. in the case of the CDR-based index the holidays were normalised by the Sunday in the reference week even when they fell on weekdays. In conclusion, the two methods are in good agreement, and both are able to follow the changes in the movement of the population at the scales relevant for the decision making related to the fight against COVID-19.

**Figure 4.**
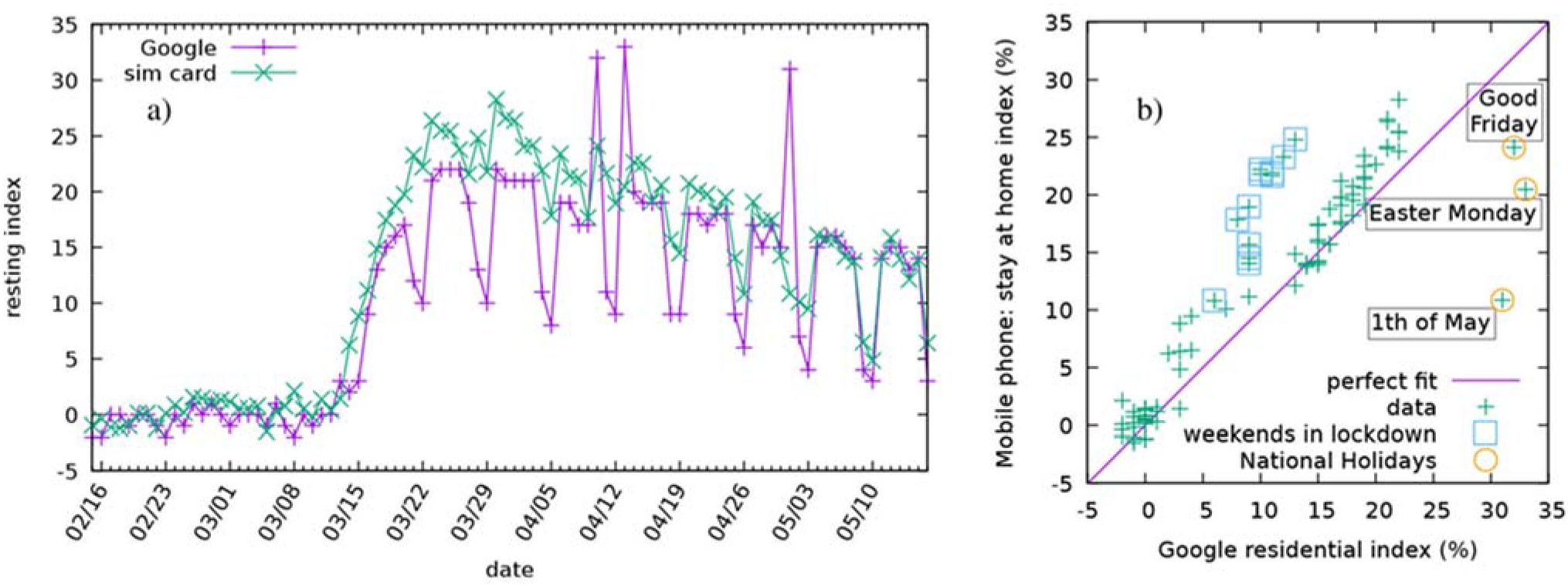
Comparison of two methods for measuring the relative change from the baseline of “staying-at-home”. Left **(a)**: The residential index of Google (purple) and the CDR based stay-at-home index (green) plotted as a time series. Right **(b)**: Scatter plot of the daily stay-at-home index as a function of the daily residential index of Google. Perfect fit would correspond to a set of points falling onto the diagonal line shown in purple. The weekends during the curfew seem to deviate from this to the left, whereas outliers to the right are corresponding to the national holidays.

## Discussion

The idea to use mobile phone CDR to track population movement in the case of emergencies is not new, and its use has been reported during this COVID-19 pandemic.^21^ Nevertheless, to our knowledge, no detailed description of their application and utility has been published to date.

We have shown that the mobility and staying-at-home indices developed using CDR data give results that are similar, but not identical, to those with other applications that record population movement on the basis of high resolution smartphone capabilities. Nevertheless, both approaches have strengths and limitations. The advantage of smartphone-generated, satellite-based location data, such as that collected by Goggle, is that the temporal and spatial resolution is extremely high for any individual,^38^ which is impossible to achieve with CDR. In addition, by linking its mobility data to the vast amount of customer data owned by Google and similar companies it is possible to study mobility by characteristics of the phone owner, including by socio-economic position or employment, whose feasibility is limited using CDR only. However, a serious disadvantage is that only some users are included. Google see only Android users, and among them only those, whose location history is turned on. In general, any smartphone-based location tracking solution is blind to individuals, who do not have a smartphone, or not willing to share their location. In contrast, by combining the data from all mobile phone operators, generated automatically, as part of the routine administration of calls, the CDR-based mobility index captures all active mobile phone users. This property makes its implementation relatively easy and inexpensive, and can be especially important in settings where early versions of phones are still widely used. In Hungary, out of roughly 8.6 million mobile phone users, 5.3 million have smart phones, further broken down to 4.5 million Android users and 0.8 million using some other operating system.^39^ Therefore, the mobility index using CDR benefits from having data from almost twice as many users as Google. While this may not seem to be a major advantage at the national level, it can make a substantial difference in smaller geographical areas, where smartphone penetration is small, or where a significant part of the population deny access to their location data. Voluntary compliance is the Achilles-heel of any smartphone-based location tracking application, and while mandating its use can work for small scale, targeted epidemiological interventions, it would be a serious infringement on civil liberties if it was enforced for whole populations.

The main limitation of the CDR-based method is that the accuracy of localisation depends on the use of the phone. Thus, it will only register when a call is made, an SMS is sent, or data accessed. Moreover, if a user is starting a long phone call in one settlement and continues to talk during a trip to another distant one, the CDR-based location method will report the client as if they remain at the starting position. So, no movement is detected by the method, when the location of the phone did change. However, if the same client is making a call using data via an app, such as WhatsApp, the trip will be tracked accurately because apps usually divide their data flow into shorter sessions. Therefore, a new session event, with the changed location information, is generated frequently, allowing geolocation at high resolution.

In summary, as shown in Table 1, the CDR-based tracking is superior to the smartphone-based tracking in that the former is automatically generated for all users, while the latter is only for those, who have smartphones and give their permission for location tracking. On the other hand, the CDR-based method depends on the actual use of the device, which makes the tracking of less frequent phone users less accurate. This limitation could be overcome by using cell registration data, which is feasible, but they are more difficult to obtain and process. Another limitation is that the socioeconomic characteristics could not be linked to the basic CDR event records. Such analysis is feasible only with more detailed device registry data. The fact that the CDR-based method, as used by the government, utilises anonymous, aggregated data provided by independent operators, makes the identification of individual users impossible. On the other hand, it is rather a strength than a weakness of the methodology in that no personal data protection and privacy questions arise. The aggregation and anonymization of the data were guaranteed by the mobile network operators, since they had to hide details of CDR records not only for the protection of their customers, but to protect details of their business strategies too.

**Table 1.** Comparison of CDR-based tracking as opposed to smartphone-based tracking of mass population movement.

|  | **CDR-based method** | **Smartphone-based method** |
| --- | --- | --- |
| **Location information** | Cellular base station (mast) | Satellite and cellular base station |
| **Tracking accuracy** |  |  |
| **Location of a person** | Lower (around one hundred metres to kilometres), depends on mast density and environment<br>Depends on the use of the phone | Higher (few metres with satellite positioning) |
| **Population coverage** | Higher | Lower |
| - people without mobile phones | Not covered | Not covered |
| - people with mobile phones turned off | Not covered | Not covered |
| - people with ordinary mobile phones | Covered | Not covered |
| - people with smartphones location function turned off | Covered | Not covered |
| - people with smartphones location function turned on | Covered | Covered |
| **Analysis of mobility by personal characteristics** | Very limited | Yes |
| **Feasibility** | Easier and less expensive | More expensive |
| **Personal data protection and privacy** | Ensured with anonymous and aggregated data<br>Does not require approval from mobile phone users (approval is given when one registers for the service) | Requires approval from smartphone users |
| **When should be used?** | Better option for monitoring mass population movement during large scale restrictions country-wide (hammer phase), or regional (dance phase) | Better to track individuals or smaller population groups for the purpose of monitoring targeted interventions, such as contact tracing or quarantine monitoring |

The experience in Hungary has shown that CDR-based mobility and staying-at-home indices provide a means to monitor the effectiveness of restrictions on mobility and to pinpoint problematic local geographical areas where further measures are warranted. Swift implementation of strict measures to reduce the frequency of movement seems to be paying off as seen by the continuing low incidence of infection. In the early phase of the COVID-19 outbreak, the Hungarian government was able to interrupt the exponential growth of new cases, keep the curve flat and keep the number of COVID-19 deaths per 1 million population well below the average of the EU-27 and the UK. The next test of the methodology will be the exit phase, where success depends on the accuracy and timeliness of monitoring, as well as the agility of contact tracing, should a local flare up occur. While the CDR-based method seems to be a better option to monitor mass population movement during large scale restrictions, when there is no need to pinpoint the location of individuals with high precision but it is very important to see as many members of the population as possible, the smartphone-based method should work better to track individuals or smaller population groups for the purpose of monitoring targeted interventions, such as contact tracing or quarantine monitoring, where high precision is the main determinant of effectiveness. In this respect, the two methods are rather complementary in supporting pandemic responses.

## Conclusions

We report how integrated CDR data from three major European telecommunication companies covering almost all the mobile phone market in Hungary have been used to monitor mobility. The algorithms automatically process these huge routine databases into information easily interpreted by high level decision makers. The process has been made routine and sufficiently rapid to support swift decision making. We are convinced, therefore, that it can be easily adapted by other countries, should they wish to apply it in the management of the COVID-19 outbreak. The Digital Health and Data Utilisation Team is happy to share it upon request to support the fight against the COVID-19 pandemic all over the world.

## Data Availability

The data available at Harvard Dataverse as Hun Covid Mobile Dataset.

https://doi.org/10.7910/DVN/IPJULW

## Acknowledgements and funding information

Semmelweis University is a government funded public institution. A specific grant supported the operation of the DHDUT from the National Research, Development and Innovation Fund (grant no. 2020-2.1.1-ED-2020-00003), and another grant of the Hungarian National Research, Development and Innovation Office also supported the research (grant no. K 128780). The research was partially financed by the Research Excellence Programme of the Ministry for Innovation and Technology in Hungary, within the framework of the Digital Biomarker thematic programme of the Semmelweis University.

MT, TM and Vodafone Hungary provided the datasets and their expertise free of charge.

The authors thank for the fruitful discussions with Gergely Palla and the comments and suggestions of Josep Figueras.

## Author Contributions

M.Sz. conceived and led the study including the setup of cooperation between the independent providers, participated in the implementation and analysis, and contributed to the writing of the paper, P.P. defined the GDPR compatible aggregation method, I.S. and T.J. implemented the calculations and compiled the reporting system for the decision makers, P.P., I.S., T.J., T.P. and M.M. participated in the implementation and the analysis, and contributed to the writing of the paper, P.G. participated in the implementation and the analysis of the results, put together the concept and wrote the first draft of the paper, contributed to the subsequent revisions and prepared the final version of the paper. M.T., T.M., Á.S., J.Sz., Á.T. collected, preprocessed and provided the anonymous dataset respectively. All authors reviewed and accepted the manuscript.

## References

1. Bi Q, Wu Y, Mei S, et al. Epidemiology and transmission of COVID-19 in 391 cases and 1286 of their close contacts in Shenzhen, China: a retrospective cohort study. Lancet Infect Dis 2020 doi: 10.1016/S1473-3099(20)30287-5 [published Online First: 2020/05/01]

2. Chinazzi M, Davis JT, Ajelli M, et al. The effect of travel restrictions on the spread of the 2019 novel coronavirus (COVID-19) outbreak. Science 2020;368(6489): 395-400. doi: 10.1126/science.aba9757 [published Online First: 2020/03/08]

3. Maffioli EM. How Is the World Responding to the 2019 Coronavirus Disease Compared with the 2014 West African Ebola Epidemic? The Importance of China as a Player in the Global Economy. Am J Trop Med Hyg 2020 doi: 10.4269/ajtmh.20-0135 [published Online First: 2020/03/13]

4. Nature. Coronavirus updates: the first three months as it happened. http://dx.doi.org/10.1038/d41586-020-00154-w 2020.

5. WHO Regional Office for Europe, European Commission, European Observatory on Health Systems and Policies. Covid-19 Health System Response Monitor web page 2020 [Available from: https://www.covid19healthsystem.org/mainpage.aspx accessed 21/05 2020.

6. World Health Organization. Novel Coronavirus - China. http://www.who.int/csr/don/12-january-2020-novel-coronavirus-china/en/, 2020.

7. World Health Organization. Novel Coronavirus - Situation Report 87. http://www.who.int/docs/default-source/coronaviruse/situation-reports/20200416-sitrep-87-covid-19.pdf?sfvrsn=9523115a_2, 2020.

8. The Center for Systems Science and Engineering. Coronavirus COVID-19 Global Cases. http://www.arcgis.com/apps/opsdashboard/index.html\#/bda7594740fd40299423467b48e9ecf6: Johns Hopkins University, 2020.

9. Anderson RM, Heesterbeek H, Klinkenberg D, et al. How will country-based mitigation measures influence the course of the COVID-19 epidemic? Lancet 2020; 395: 931–34. doi: https://doi.org/10.1016/S0140-6736(20)30567-5

10. McKibbin W, Fernando R. The Global Macroeconomic Impacts of COVID-19: Seven Scenarios. CAMA Working Paper No 19/2020 2020 doi: http://dx.doi.org/10.2139/ssrn.3547729

11. Nicola M, Alsafi Z, Sohrabi C, et al. The Socio-Economic Implications of the Coronavirus and COVID-19 Pandemic: A Review. Int J Surg 2020 doi: 10.1016/j.ijsu.2020.04.018 [published Online First: 2020/04/20]

12. Sohrabi C, Alsafi Z, O’Neill N, et al. World Health Organization declares global emergency: A review of the 2019 novel coronavirus (COVID-19). Int J Surg 2020; 76: 71-76. doi: 10.1016/j.ijsu.2020.02.034 [published Online First: 2020/03/01]

13. McKee M, Stuckler D. If the world fails to protect the economy, COVID-19 will damage health not just now but also in the future. Nat Med 2020 doi: 10.1038/s41591-020-0863-y [published Online First: 2020/04/11]

14. McKee M. Learning from success: how has Hungary responded to the COVID pandemic? GeroScience 2020: 1–3. doi: 10.1007/s11357-020-00240-x [published Online First: 2020/07/29]

15. Gaal P, Szerencses V, Velkey Z, et al. Covid-19 Health System Response Monitor: Hungary web page 2020 [Available from: https://www.covid19healthsystem.org/countries/hungary/countrypage.aspx accessed 21/05 2020.

16. Balzotti C, Bragagnini A, Briani M, et al. Understanding Human Mobility Flows from Aggregated Mobile Phone Data⁎⁎This work was supported by funding from project MIE - Mobilità Intelligente Ecosostenibile (CTN01_00034_594122), Cluster “Tec-nologie per le Smart Communities”. IFAC-PapersOnLine 2018; 51(9): 25–30. doi: https://doi.org/10.1016/j.ifacol.2018.07.005

17. Blondel VD, Decuyper A, Krings G. A survey of results on mobile phone datasets analysis. EPJ Data Science 2015; 4(1): 10. doi: 10.1140/epjds/s13688-015-0046-0

18. Eagle N. Mobile Phones as Social Sensors. In: Hesse-Bieber SN, ed. The Handbook of Emergent Technologies in Social Research. New York: Oxford University Press 2011: 492–521.

19. Palla G, Barabási A-L, Vicsek T. Quantifying social group evolution. Nature 2007;446(7136): 664–67. doi: 10.1038/nature05670

20. Google Inc. Google Maps Timeline web page: Google Inc; 2020 [Available from: https://support.google.com/maps/answer/6258979?co=GENIE.Platform%3DDesktop&hl=en accessed 01/06 2020.

21. Oliver N, Lepri B, Sterly H, et al. Mobile phone data for informing public health actions across the COVID-19 pandemic life cycle. Science Advances 2020: 1–10. doi: 10.1126/sciadv.abc0764 [published Online First: 27/04/2020]

22. Kamel Boulos MN, Geraghty EM. Geographical tracking and mapping of coronavirus disease COVID-19/severe acute respiratory syndrome coronavirus 2 (SARS-CoV-2) epidemic and associated events around the world: how 21st century GIS technologies are supporting the global fight against outbreaks and epidemics. Int J Health Geogr 2020; 19(1): 8. doi: 10.1186/s12942-020-00202-8 [published Online First: 2020/03/13]

23. Buchwald E. What we can learn from South Korea and Singapore’s efforts to stop coronavirus (besides wearing face masks) web page: MarketWatch Inc.; 2020 [updated 6 April 2020. Available from: https://www.marketwatch.com/story/what-we-can-learn-from-south-korea-and-singapores-efforts-to-stop-coronavirus-in-addition-to-wearing-face-masks-2020-03-31 accessed 30/04 2020.

24. Ferretti L, Wymant C, Kendall M, et al. Quantifying SARS-CoV-2 transmission suggests epidemic control with digital contact tracing. Science 2020: eabb6936. doi: 10.1126/science.abb6936

25. Armocida B, Formenti B, Ussai S, et al. The Italian health system and the COVID-19 challenge. Lancet Public Health 2020 doi: 10.1016/S2468-2667(20)30074-8 [published Online First: 2020/03/30]

26. Karnitschnig M. The incompetence pandemic: The first victim of the coronavirus? Leadership.: Politico LLC; 2020 [updated 16 March 2020. Available from: https://www.politico.com/news/2020/03/16/coronavirus-pandemic-leadership-131540 accessed 30 April 2020.

27. Government of Hungary. Decree No. 40/2020. (III.11.) Korm. on the Declaration of a State of Emergency. Hungarian Gazette, 2020;2020(39): 1354.

28. Government of Hungary. Decree No. 41/2020. (III.11.) Korm. on the the Measures to be Taken during the State of Danger Declared for the Prevention of the Human Epidemic Endangering Life and Property and Causing Massive Disease Outbreaks, for the elimination of its Consequences, and for the Protection of the Health and Lives of Hungarian Citizens. Hungarian Gazette, 2020;2020(40): 1356–58.

29. Government of Hungary. Decree No. 46/2020. (III.16.) Korm. on the Measures to be Taken during the State of Danger Declared for the Prevention of the Human Epidemic Endangering Life and Property and Causing Massive Disease Outbreaks, for the elimination of its Consequences, and for the Protection of the Health and Lives of Hungarian Citizens (III). Hungarian Gazette. 2020;2020(45): 1444–46.

30. Government of Hungary. Decree No. 71/2020. (III.27.) Korm. on Restricting Movement. Hungarian Gazette, 2020;2020(56): 1626–27.

31. Government of Hungary. Resolution No. 1102/2020. (III.14.) Korm. on the Introduction of a New Work Arrengment in Public Schools and Vocational Training Institutions. Hungarian Gazette, 2020;2020(43): 1388.

32. Government of Hungary. Decree No. 118/2020. (IV.16.) Korm. on the Authorisation of Local Governments to Implement the Restriction of Movement regarding the Weekend. Hungarian Gazette, 2020;2020(77): 2026.

33. Government of Hungary. Article 6, point a) of Decree No. 45/2020. (III.14.) Korm. on the Measures to be Taken during the State of Danger Declared for the Prevention of the Human Epidemic Endangering Life and Property and Causing Massive Disease Outbreaks, for the elimination of its Consequences, and for the Protection of the Health and Lives of Hungarian Citizens (II). Hungarian Gazette, 2020;2020(43): 1382-83.

34. Government of Hungary. Article 14 of Decree No. 46/2020. (III.16.) Korm. on the Measures to be Taken during the State of Danger Declared for the Prevention of the Human Epidemic Endangering Life and Property and Causing Massive Disease Outbreaks, for the elimination of its Consequences, and for the Protection of the Health and Lives of Hungarian Citizens (III). Hungarian Gazette, 2020;2020(45): 1444–46.

35. Government of Hungary. Article 10 of Decree No. 46/2020. (III.16.) Korm. on the Measures to be Taken during the State of Danger Declared for the Prevention of the Human Epidemic Endangering Life and Property and Causing Massive Disease Outbreaks, for the elimination of its Consequences, and for the Protection of the Health and Lives of Hungarian Citizens (III). Hungarian Gazette, 2020;2020(45): 1444–46.

36. National Media and Infocommunications Authority. A Nemzeti Média és Hírközlési Hatóság mobilpiaci jelenetése, 2015 IV. - 2019. IV. negyedév [The Mobile Market Report of the NMHH for the period of Q4 2015 to Q4 2019]. http://nmhh.hu/dokumentum/211976/NMHH_mobilpiaci_jelentes_2015Q42019Q4.pdf: NMHH, 2020: 26.

37. Hungarian Central Statistical Office. Regional Atlas – Settlements. http://www.ksh.hu/regionalatlas_settlements?lang=en: Központi Statisztikai Hivatal, 2020.

38. Google Inc. COVID-19 Community Mobility Reports. https://www.google.com/covid19/mobility, 2020.

39. eNET Internet Research Consulting Ltd. 5,3 million smartphone users in Hungary. https://enet.hu/news/5-3-million-smartphone-users-in-hungary, 2019.

